# Epitope-resolved serology test differentiates the clinical outcome of COVID-19 and identifies defects in antibody response in SARS-CoV-2 variants

**DOI:** 10.1101/2021.03.16.21253716

**Authors:** Courtney Voss, Sally Esmail, Xuguang Liu, Michael J. Knauer, Suzanne Ackloo, Tomonori Kaneko, Lori Lowes, Peter Stogios, Almagul Seitova, Ashley Hutchinson, Farhad Yusifov, Tatiana Skarina, Elena Evdokimova, Peter Loppnau, Pegah Ghiabi, Taraneh Haijian, Shanshan Zhong, Husam Abdoh, Benjamin D. Hedley, Vipin Bhayana, Claudio M. Martin, Marat Slessarev, Benjamin Chin-Yee, Douglas D. Fraser, Ian Chin-Yee, Shawn S-C. Li

**Author notes:** These authors contributed equally to this work (Courtney Voss, Sally Esmail).

## Abstract

**BACKGROUND:** The role of humoral immunity in the coronavirus disease 2019 (COVID-19) is not fully understood owing, in large part, to the complexity of antibodies produced in response to the SARS-CoV-2 infection. There is a pressing need for serology tests to assess patient-specific antibody response and predict clinical outcome.

**METHODS:** Using SARS-CoV-2 proteome and peptide microarrays, we screened 146 COVID-19 patients plasma samples to identify antigens and epitopes. This enabled us to develop a master epitope array and an epitope-specific agglutination assay to gauge antibody responses systematically and with high resolution.

**RESULTS:** We identified 54 linear epitopes from the Spike (S) and Nucleocapsid (N) protein and showed that epitopes enabled higher resolution antibody profiling than protein antigens. Specifically, we found that antibody responses to the S(811-825), S(881-895) and N(156-170) epitopes negatively or positively correlated with clinical severity or patient survival. Moreover, we found that the P681H and S235F mutations associated with the coronavirus variant B.1.1.7 altered the specificity of the corresponding epitopes.

**CONCLUSIONS:** Epitope-resolved antibody testing not only offers a high-resolution alternative to conventional immunoassays to delineate the complex humoral immunity to SARS-CoV-2 and differentiate between neutralizing and non-neutralizing antibodies, it may also be used as predictor of clinical outcome. The epitope peptides can be readily modified to detect antibodies against variants in both the peptide array and latex agglutination formats.

**FUNDING:** Ontario Research Fund (ORF)-COVID-19 Rapid Research Fund, the Toronto COVID-19 Action Fund, Western University, the Lawson Health Research Institute, the London Health Sciences Foundation, and the AMOSO Innovation Fund.

## INTRODUCTION

The severe acute respiratory syndrome coronavirus-2 (SARS-CoV-2) has infected more than 100 million people worldwide since it was first identified in humans in 2019. The ensuing COVID-19 pandemic has put diagnostic testing at the forefront in the battle to stop the spread of the virus. Nucleic acid testing (NAT), which detects the virus RNA by reverse transcription polymerase chain reaction (RT-PCR), is the current gold standard for diagnosing acute infections(1). NAT has played a critical role in containing the pandemic by allowing expedient identification of infected individuals for treatment, isolation and contact-tracing. However, NAT alone cannot reveal the true prevalence of the SARS-CoV-2 infection because 20%-80% of all infections are likely asymptomatic(2-4). Therefore, a significant proportion of the population would be missed by NAT-based screening because the virus is typically cleared by the immune system in 3-4 weeks after infection or symptom onset. To complement NAT, serological assays for virus-specific antibodies have been developed(5-7). In contrast to NAT that can only detect acute infections, serology tests can identify past infections as antibodies may persist in the blood long after the virus has been cleared. The wide window of time within which antibodies may be detected, ranging from 1-2 weeks of infection when seroconversion occurs to several months after the infection is resolved, offers a unique advantage for antibody testing over NAT. Because of the high incidence of asymptomatic cases, antibody testing, when carried out in large scales, can provide valuable and accurate information about the spread of the infection at the population level and the true infection fatality rate(8, 9). Importantly, with the advent of several effective vaccines against the virus and the rapid rollout of the vaccination program around the world, priorities are being shifted from containment to monitoring the immediate and longitudinal effects of the vaccines on the immune system. This paradigm shift will undoubtedly increase the demand for antibody testing.

Numerous serological assays for SARS-CoV-2 antibodies have been developed to date, including enzyme-linked immunosorbent assays (ELISA), chemiluminescent immunoassays (CLIA) and lateral flow assays (LFAs) (1, 8). The sensitivity and specificity of different ELISA kits may vary(10), but they are generally considered sufficient for large-scale SARS-CoV-2 antibody testing. Nevertheless, the need for specialized equipment and trained personnel to perform the test and the long turn-around time makes it a challenge to use ELISA in point-of-care (POC) settings. In contrast, LFAs, which can be carried out in under 30 minutes with no equipment required, can potentially be used for POC testing. However, LFA-based tests have been shown less sensitive and specific than ELISA (6, 9, 11, 12). Besides concerns over sensitivity, specificity and POC potential, both ELISA- and LFA-based antibody testing have the following limitations. First, current tests rely on the interaction of the Spike (S) or Nucleocapsid (N) protein or a fragment/domain of either protein to capture the corresponding antibody. These assays, which provide a single measure of antibody reactivity, are not ideal for gauging the diverse antibody responses observed in the clinic. Second, protein antigen-based immunoassays such as ELISA and LFA generate a composite signal across many epitopes, including both conformational and linear epitopes, and thereby lacking the necessary specificity or resolution to differentiate between neutralizing and non-neutralizing antibodies or predict clinical outcome. Indeed, patients who are older or with severe symptoms have been shown to produce more antibodies than those who are younger or with milder symptoms(13, 14), suggesting that robust antibody responses measured by conventional means do not correlate with effective humoral immunity. Third, current serological assays are ill-suited to assess the immunological effect of coronavirus variants as numerous recombinant proteins would have to be produced. Several mutated strains have emerged recently that are believed to be more contagious than the original SARS-CoV-2 strain(15, 16). These and other variants identified to date harbor numerous missense or deletion mutations in the S or N protein encoding gene that may alter their antigenic characteristics. To effectively curb the spread of these highly contagious variants, it is of paramount importance that we develop an antibody test that can readily incorporate the emerging mutations to determine the effect of these mutations and the corresponding coronavirus variants on the immune system. Fourth, current immunoassays are generally focused on testing a specific antibody isotype. Given the distinct dynamics of IgM, IgA and IgG in response to the SARS-CoV-2 infection(17, 18), it is necessary to develop a multiplex immunoassay to gauge humoral immunity. Lastly, with the vaccine rollout across the globe, a rapid and accurate POC test is urgently needed to gauge the effectiveness of a vaccine and monitor the duration of antibody responses in large populations to provide valuable information on herd immunity.

We addressed these unmet needs in SARS-CoV-2 antibody testing using protein and peptide arrays, which led to the identification of linear epitopes that mediate the complex antibody responses observed in a group of 89 COVID-19 patients. This, in turn, allowed us to develop a “master epitope array” containing the major epitopes and use it to gauge antibody responses with greater resolution than is attainable by protein antigen-based immunoassays. We found that the antibody profiles determined by linear epitopes, but not by the S or N protein, could distinguish patients with moderate or severe diseases or with favorable or fatal outcomes. Using a peptide array recapitulating the mutations found in SARS-CoV-2 variants, we showed that certain mutations abolished binding of the corresponding epitopes to antibodies against the original strain. Furthermore, the identified epitopes enabled us to develop an epitope-dependent agglutination assay for SARS-CoV-2 antibodies. This rapid agglutination assay is not only highly accurate, but it may also be readily modified to incorporate specific epitopes, including variant epitopes, to profile the complex antibody responses in individuals.

## RESULTS

### Antibody responses to the S or N protein are not correlated with clinical outcome

To develop a comprehensive antibody test, we first employed a protein array to identify the SARS-CoV-2 antigens mediating antibody responses. Previous studies have implicated the S, N and the non-structural proteins encoded by the *ORF1ab* gene as the major antigens eliciting humoral immune response in the host(19, 20). We therefore expressed these proteins, including various different fragments or domains of S and N, in bacterial or mammalian cells. Upon purification, the recombinant virus proteins were printed on nitrocellulose coated glass slides. The resulting proteome array, featuring 16 SARS-CoV-2 proteins and human IgG as the positive control (Fig. 1A, Table S1), was probed with plasma samples from patients that tested positive or negative for SARS-CoV-2 by RT-PCR (10). The bound IgG was detected using goat anti-human IgG conjugated to the horseradish peroxidase (HRP) (Fig. S1).

**Figure 1.**
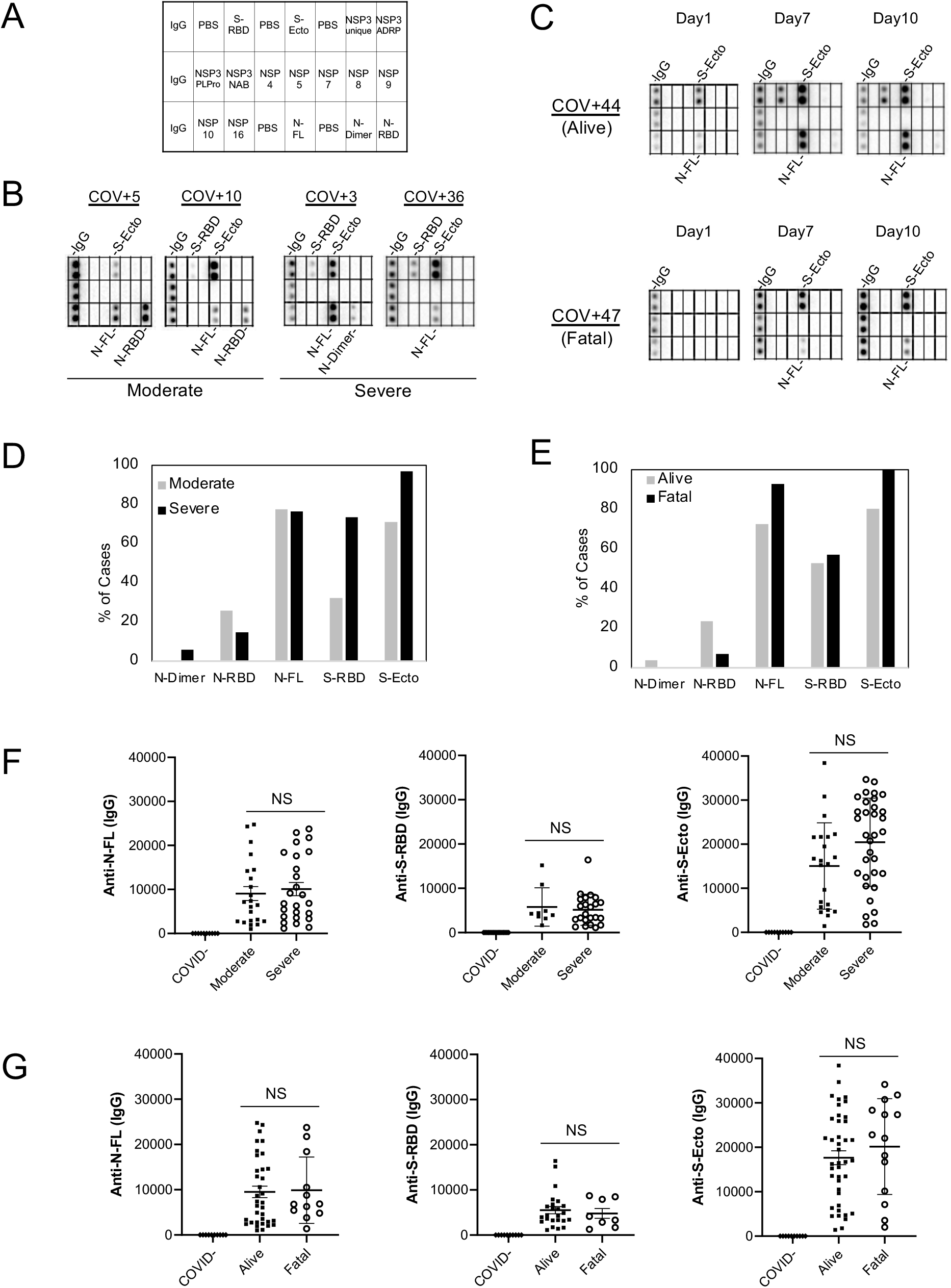
Lack of correlation between S/N antibody response and disease severity or outcome. (**A**) Layout of the proteome array. (**B**) Representative images of antibody responses for COVID-19 patients with moderate or severe disease determined using the proteome array. (**C**) Dynamic IgG antibody profiles for two patients with severe or fatal disease on days 1, 7 and 10 of ICU admission. (**D, E**) Prevalence of antibody responses to the S or N protein/domain for the indicated patient groups. (**F, G**) The intensity of antibody response to the S or N protein/domain is not correlated with disease severity (**F**) or outcome (**G**). NS, not significant; based on unpaired Student’s t-test with Welch’s correction.

We screened the proteome array and subsequent peptide arrays (*vide infra*) with 146 plasma samples from 89 hospitalized patients, including serial samples collected for some patients on different days after diagnosis. The patients were divided into two groups with severe (i.e., requiring intensive care) or moderate (i.e., no intensive care required) disease. The same patient cohort was also classified according to clinical outcome into the “alive” or “fatal” group, with the former comprising those who survived the infection and the latter who ultimately succumbed to the disease. As shown in Fig. 1B, both the moderate and severe groups showed IgG responses to the Spike (including the ectodomain S-Ecto and the receptor binding domain S-RBD) and the nucleocapsid protein (including the RNA-binding domain N-RBD and the dimerization domain N-Dimer). In contrast, no significant IgG binding signal was detected for the NSP proteins (Fig. 1A-C; Fig. S2). These results are consistent with previous findings by others that Spike and nucleocapsid are the main antigenic proteins in SARS-CoV-2(19-23). For the ICU patients with serial plasma samples, we found that the S/N-specific IgG signals increased from day 1 (of ICU admission) to days 7 and 10 for both the alive and the fatal groups (Fig. 1C). This indicates that humoral immune responses became more robust with time in these patients regardless of outcome.

Overall, we found that all seroconverted patients showed IgG responses to either the S or N protein or both. A greater percentage of the severe patient group had antibodies specific for S-RBD or S-Ecto than those with moderate conditions. In contrast, the difference in N-specific IgG signal was small between the two groups (Fig. 1D). Compared to the group that survived, the fatality group more frequently exhibited S- or N-specific antibodies (Fig. 1E), suggesting once again that a robust antibody response does not necessarily translate to a favorable outcome. In corroboration of this assertion, we found no correlation between the strength of S- or N-specific IgG signal and disease severity or outcome (Fig. 1F-G). Taken together, the proteome array screen data demonstrate that the S- or N-antibody response is not a sensitive barometer of COVID-19 clinical severity or outcome.

### Systematic identification of linear epitopes by peptide microarrays

Antibody specificity is determined by epitopes on the protein antigen, including both linear and conformational epitopes(23). Because linear epitopes are small peptides (5-20 residues), they may be identified by screening peptides generated by chemical or genetic means(19, 20, 22). To identify the linear epitopes mediating the SARS-CoV-2 antibody responses, we synthesized peptides representing the candidate epitopes reported in the literature (up to October 2020)(24, 25) and printed the peptides on a nitrocellulose coated glass slide. The resulting peptide array, containing 89 reported epitopes for the S, N, and M (membrane) proteins (Fig. 2A), was probed with patient plasma samples. Intriguingly, we were only able to detect <50% of the reported epitopes in our peptide array screens (Fig. S3; Table S2). While the large discrepancy might be attributed, in part, to the different techniques used for assaying the epitope-antibody interaction, it prompted us to redefine the epitopes using the peptide array approach. To this end, we created a peptide microarray to represent the complete S and N protein sequences. The resulting “peptide-walking” array contained 333 tiled 15-mer peptides with 5-residue overlap between two consecutive peptides (Fig. 2A).

**Figure 2.**
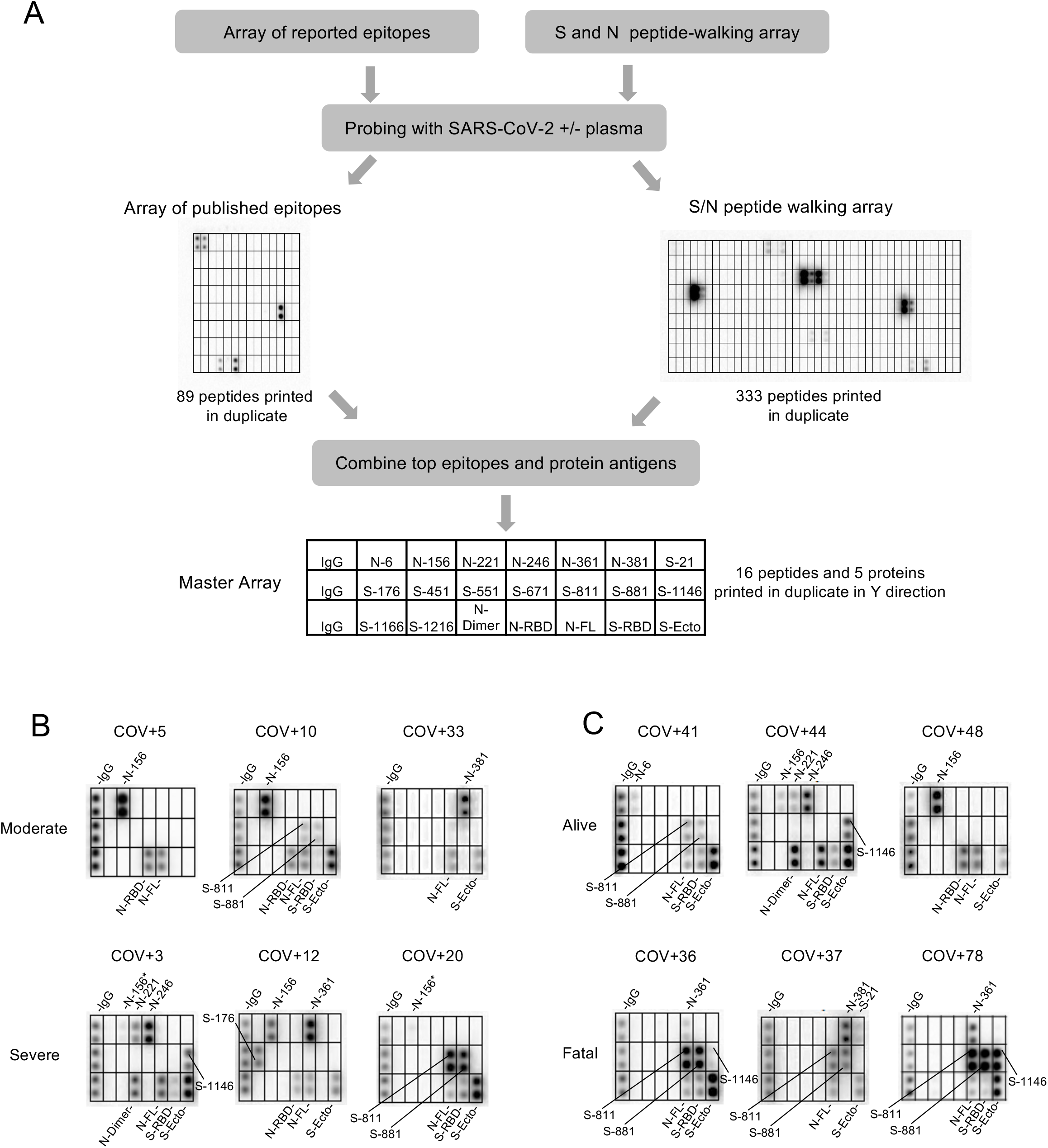
Identification of SARS-CoV-2 epitopes and epitope-resolved antibody profiling. (**A**) Workflow for identifying antigenic epitopes by peptide arrays and the layout of a master array for SARS-CoV-2 antibody profiling. (**B, C**) Representative images of epitope-resolved antibody profiles for the different groups of COVID-19 patients.

We screened the peptide microarray with 15 patient plasma samples, including 14 COVID-19 patient samples and 1 SARS-CoV-2^-^ control (Fig. S4). This led to the identification of 54 potential epitopes from the S and N protein (Table 1). While the majority of the candidate epitopes are likely minor ones based on the weak IgG-binding signals, some produced strong signals (Fig. S4, Table 1), suggesting that they may be major epitopes mediating the S or N antibody response. To profile antibody response in a systematic manner, we generated a “master epitope array” containing 16 major epitopes selected based on the corresponding IgG signal strength from the peptide-walking array screen. The master array also contained S and N protein antigens as controls (Fig. 2A; Table 1; Fig. S5).

**Table 1.**
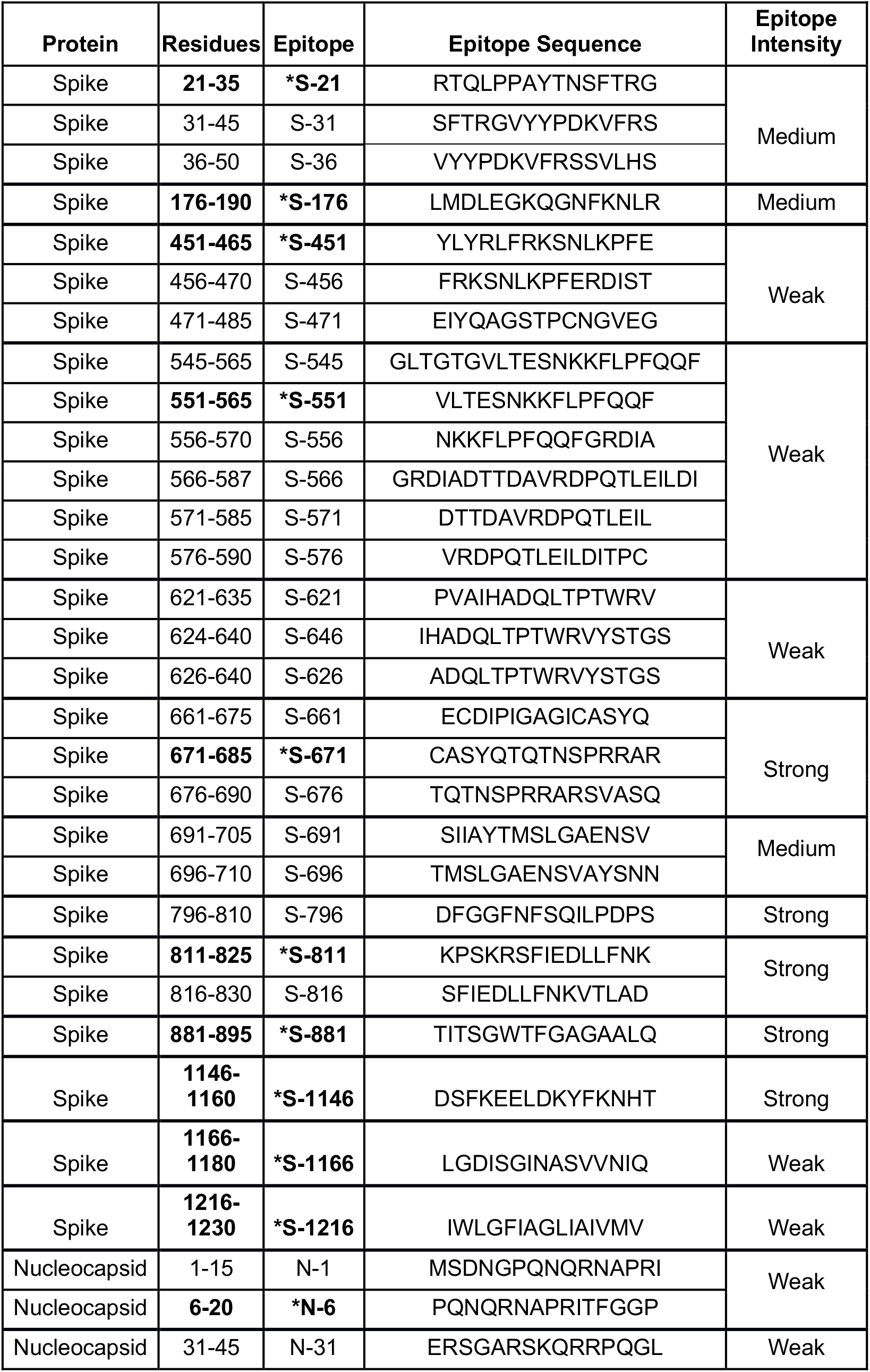
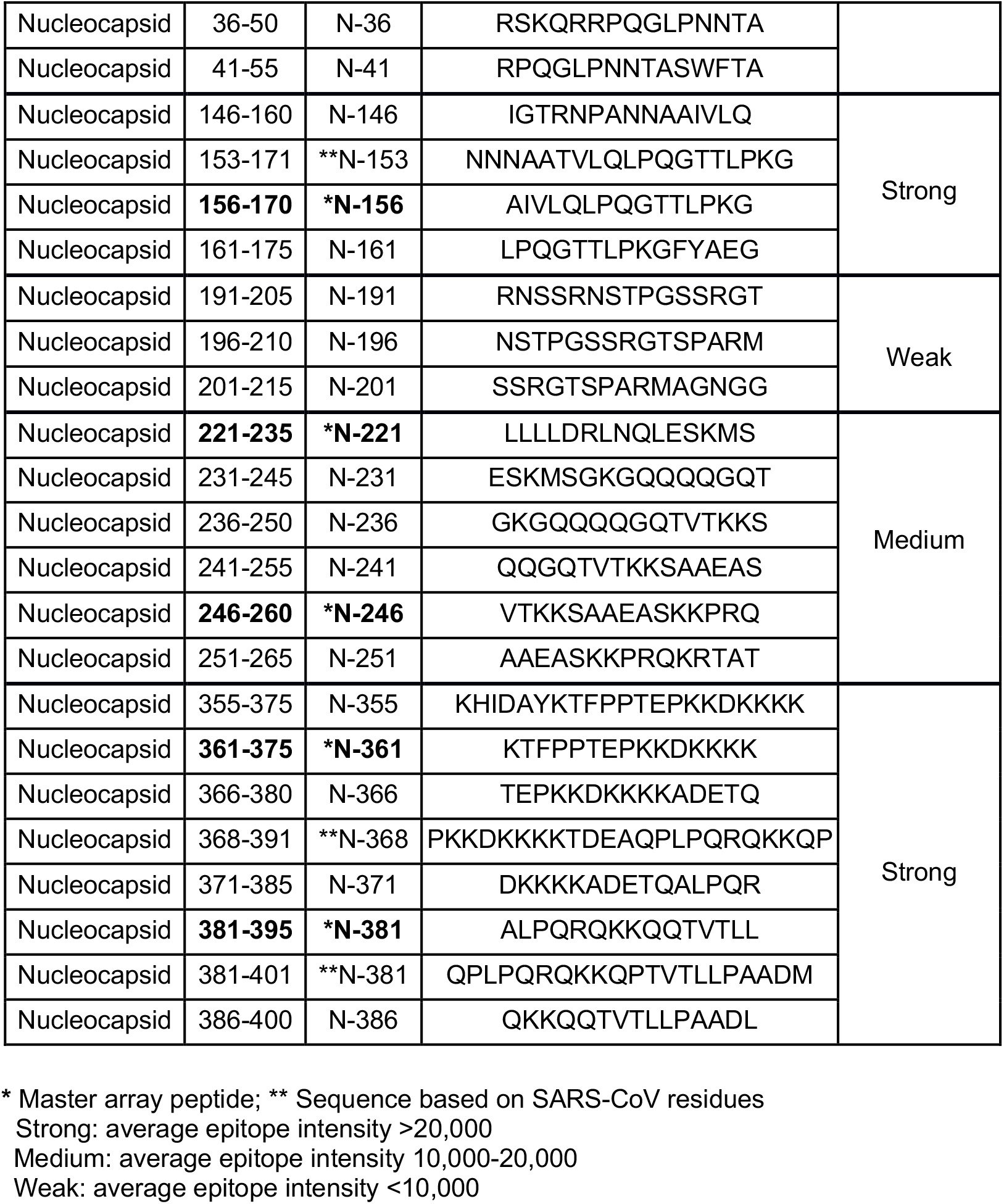
Spike and Nucleocapsid epitopes identified and characterized in this study.

### Epitope-resolved antibody profiling distinguishes COVID-19 cases based on severity or outcome

Using the master array, we screened plasma samples from the 89 COVID-19 patients and 9 SARS-CoV-2^-^ control subjects (Fig. 2 B-C; Fig. S6). We found that the plasma from ICU (severe) group recognized significantly more epitopes than the non-ICU (moderate) group (Fig. 3A). Certain epitopes, including S-811, S-881, N-6 and N-361, were detected more frequently in the severe cases than in the moderate cases whereas other epitopes, including S-451 and N-156, showed the opposite trend (Fig. 3B). By comparison, the number of IgG-binding epitopes were not significantly different between patients who survived or succumbed to the disease even though the latter group, in general, tended to have antibodies reactive to more epitopes (Fig. 3C). Nevertheless, antibodies to the S-811, S-881 and N-361 epitopes were found enriched in the fatality group whereas antibodies to N-6, S-451, S-551 and S-671 were detected only in the survivor group (Fig. 2D).

**Figure 3.**
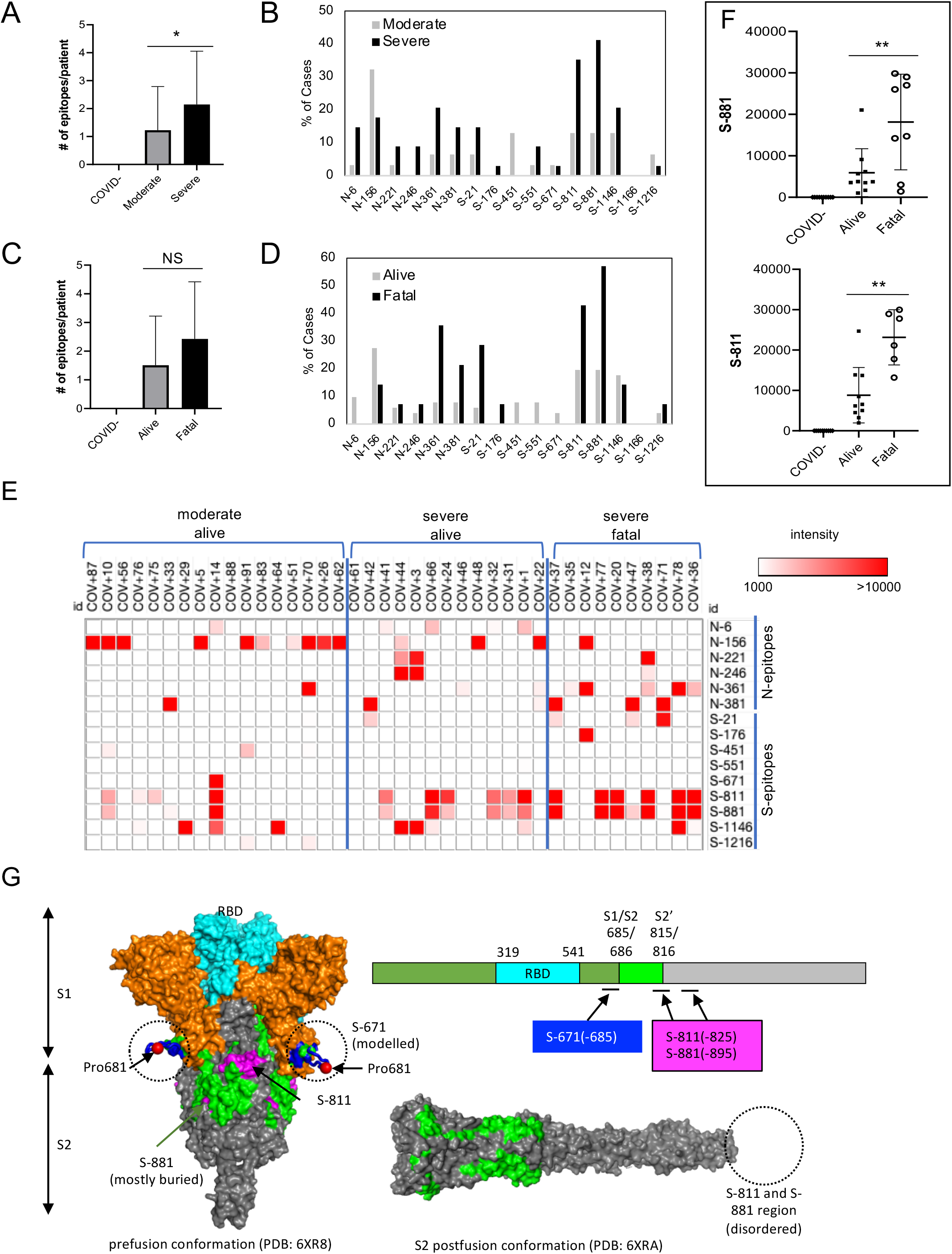
Epitope-specific antibody response distinguish COVID-19 patients with disparate disease severity and outcome. (**A**) Antibodies from patients with severe disease (n=34) recognized significantly more epitopes than those with moderate conditions (n=31). (**B**) Distribution of epitopes in moderate vs. severe cases. (**C**) Number of epitopes/patient in the alive (n=51) vs. fatal (n=14) groups. **(D)** Distribution of epitopes in alive vs. fatal cases. (**E**) Heatmap representation of epitope-specific antibodies detected by the master array. Note that only cases with at least one epitope producing IgG binding signal greater than 1000 were included in the heatmap. (**F**) Fatal cases showed significantly stronger antibody responses for the S-811 and S-881 epitopes. *, p<0.05; **, p<0.002; NS, not significant; unpaired Student’s t-test with Welch’s correction. (**G**) Structure models to show location of the critical epitopes on the Spike protein. The epitopes S-671, S-811 and S-881 are shown on the domain structure diagram of Spike as well as its prefusion (left) and post-fusion (right) conformation. The S protein has two cleavage sites, S1/S2 and S2’. The S-671 epitope is located at the C-terminus of S1, and it is disordered in the prefusion cryo-EM structure (left panel: PDB 6XR8). A homology model from the SWISS-MODEL repository was employed to draw an S-671 epitope model in the left panel (colored blue), without cleavage at S1/S2. The Pro681 site is shown with a red sphere. The S2’ cleavage site is located on the S-811 epitope. The S-881 epitope is buried and inaccessible in the prefusion state. However, the region (fusion peptide or FP) targets the host cell membrane and is fully disordered in the post-fusion conformation (right panel: PDB 6XRA, residues 771-911 are disordered). The S1 region is colored orange, except for RBD in cyan. The region between S1/S2 cleavage and S2’ cleavage sites is in green. The S-811 and S-881 epitopes are colored magenta in the prefusion conformation.

In addition to epitope frequency, the intensity of IgG-binding signals to certain epitopes were found correlated positively or negatively with clinical severity or outcome. In general, we found that moderate cases tended to have stronger antibody responses to N-156 whereas more robust antibody responses against the S-811 and S-881 epitopes were observed for the severe cases (Fig. 3E). Indeed, COV+14 was the only moderate case among the tested cases with strong S-811 and S-881 antibodies which, intriguingly also featured a robust S-671 antibody response. Overall, the patients with fatal disease were characterized with significantly stronger S-811- or S-881-specific antibodies than those who survived the infection (Fig. 3F). This indicates that antibody responses to these epitopes are detrimental to COVID-19 disease progression. The S-811 and S-881 epitopes are located in a region of the spike protein buried in the prefusion conformation, which, however, becomes disordered and exposed following virus fusion with the host cell membrane (Fig. 3G). Therefore, it is likely that the production of antibodies specific for the S-811- or S-881 epitopes coincides with the state of the coronavirus undergoing active host cell infection. In contrast, the S-671 epitope, mutated in the UK variant B.1.1.7, is located at the S1/S2 cleavage site critical for virus infection(26) (Fig. 3G).

### Mutations found in SARS-CoV-2 variants alter epitope specificity

Numerous mutations have been identified in SARS-CoV-2 variants, the vast majority of which occur on the spike protein which plays a critical role in host cell infection and immune response. The recent emergence of several variants in the United Kingdom (UK), South African (SA) and Brazil, which have been shown to be more contagious than the original strain, has raised concerns over the efficacy of mRNA vaccines that are used to produce the wild-type Spike protein in the recipient. We investigated this possibility using peptides representing 28 major S or N missense mutations or deletions identified to date, including those found in the UK variant B.1.1.7., the SA variant 501.V2 and mutations shown to alter antibody binding in a previous study(27) (Table 2). A peptide array containing the mutated epitopes and the matching counterparts in the original SARS-CoV-2 strain was created and screened with patient plasma (Fig. 4A). Because only a few mutations reside within the identified epitopes (Table 2), the mutated epitope screen was focused on plasma samples that showed robust antibody responses to the corresponding wild-type epitopes on the master array (Fig. 2B). Intriguingly, we found that the mutations either reduced or completely abolished IgG binding for the corresponding epitopes. For example, substitution of the S235 residue with a Phe in the N-221 epitope, a mutation found in the B.1.1.7 variant, eliminated IgG binding. Similarly, S-671 was identified as a major epitope in the COV+14 patient by the master array. The introduction of the P681H mutation, found in the Spike protein of the UK variant, into the S-671 peptide, completely abolished antibody binding. To confirm this finding, we synthesized another version of the S-671 epitope in which the P681 residue and the P681H mutation were placed in the center of the corresponding peptides and printed both versions of the original and mutant peptides in incremental concentrations in an array. This peptide gradient array was then probed with the COV+14 plasma collected on days 1, 2 and 3 of hospitalization. While the original epitopes exhibited increased IgG-binding with time, the P681H-containing epitope did not show detectable antibody binding signal for the same samples. These data indicate that the P681H mutation altered the specificity of the corresponding epitope (S-671) and rendered it unrecognizable by antibodies against the original coronavirus (as the plasma sample was collected prior to the emergence of the B.1.1.7 variant).

**Table 2.**
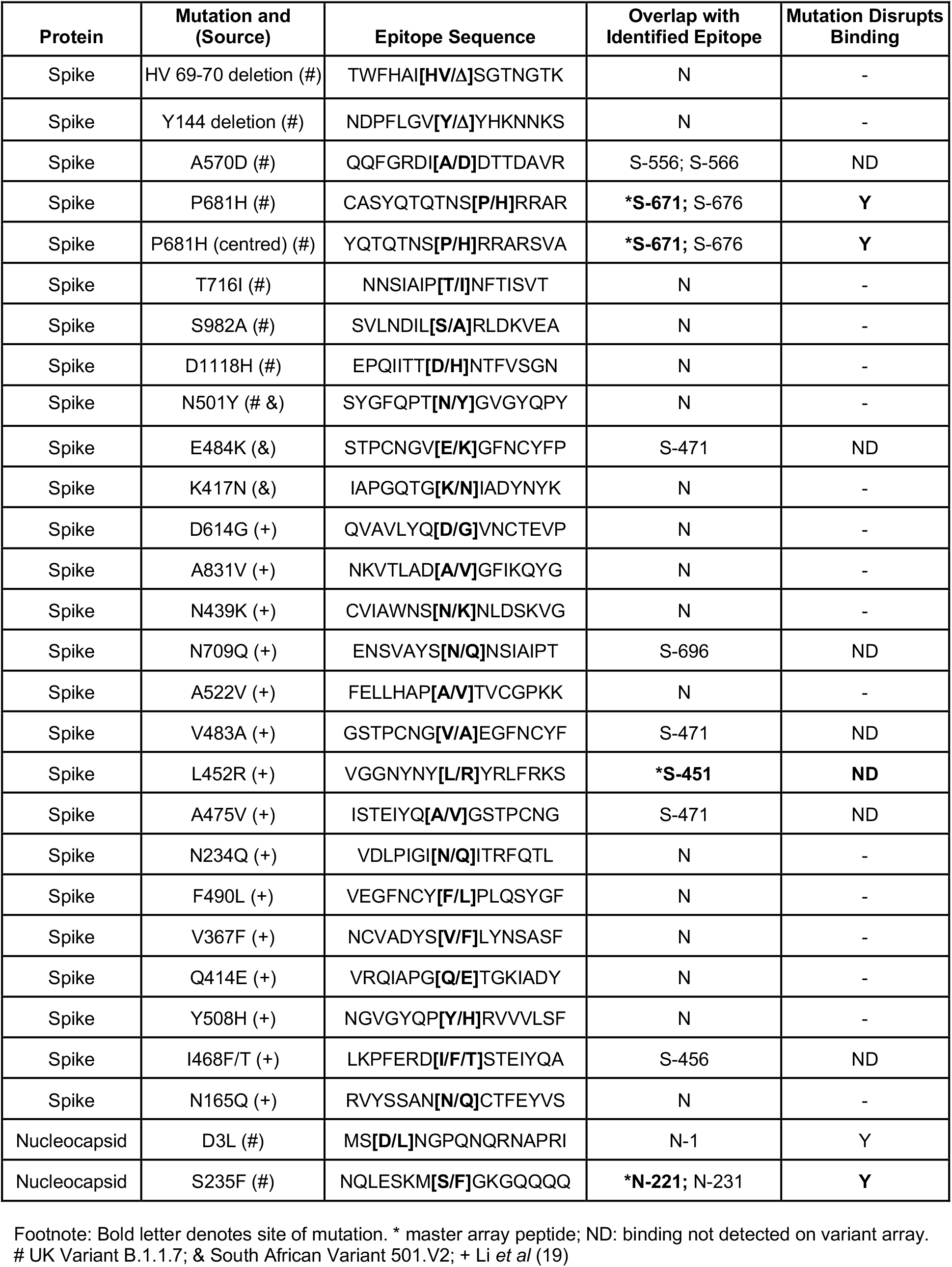
A list of S and N mutations examined by the variant epitope peptide array.

**Figure 4.**
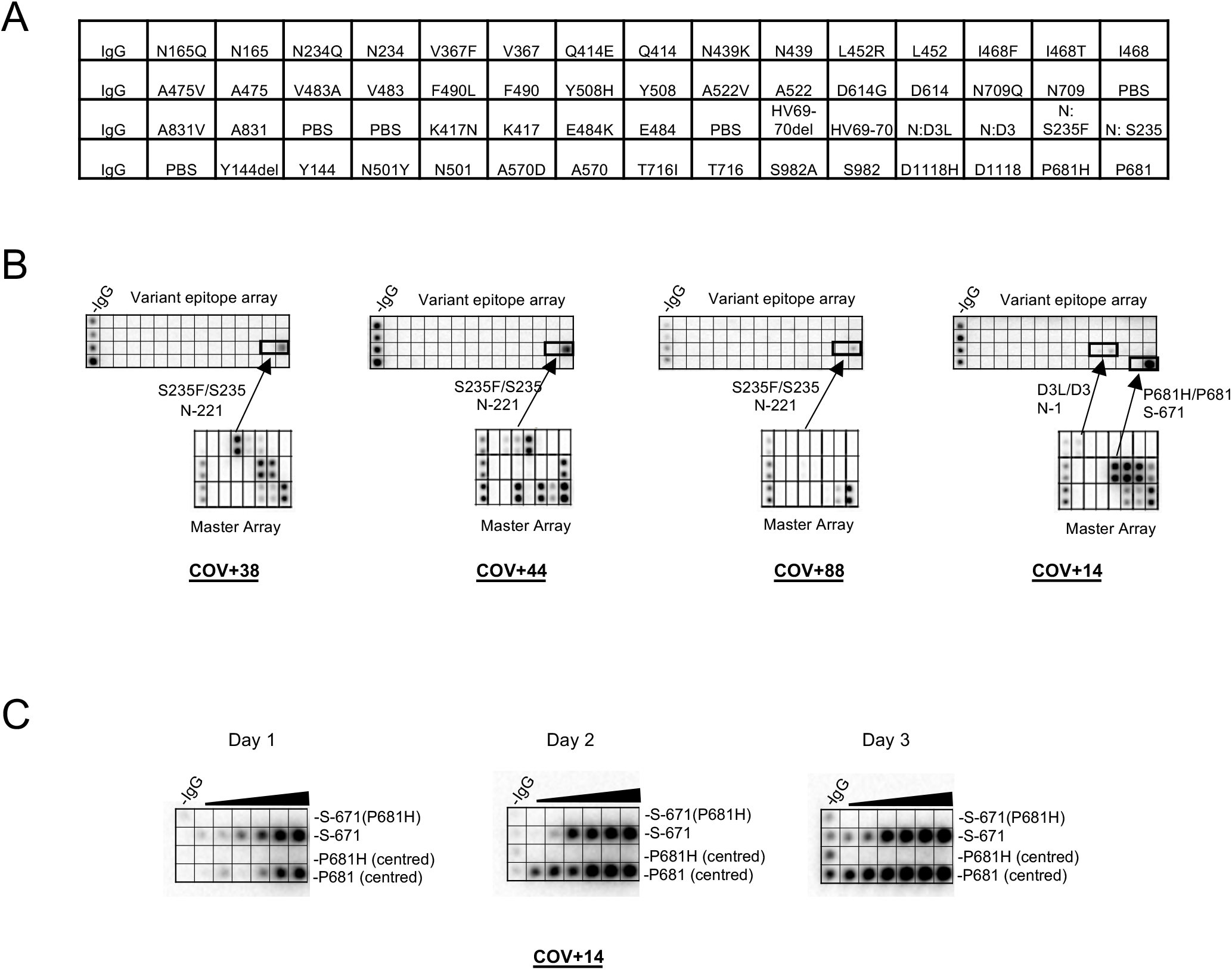
SARS-CoV-2 variants feature mutated epitopes not recognized by antibodies specific for the corresponding wt epitopes. (**A**) Layout of a SARS-CoV-2 variant epitope array. (**B**) Examples of COVID-19 cases that showed distinct IgG responses to the mutated and wt epitopes (boxed). (**C**) Dilution series of P681/P681H-containing epitopes demonstrating the loss of binding for the mutant epitopes.

### A rapid agglutination assay to gauge epitope-specific antibody response

While the epitope peptide array may be used to determine antibody specificity in a systematic manner, it is not suitable for POC testing. Nevertheless, the identification of specific epitopes that are either common to the COVID-19 patients examined or unique to groups with distinct clinical severity or outcome prompted us to develop a rapid test based on these epitopes to complement the peptide array assay. Inspired by the principle of antibody-dependent red blood cell agglutination(28), we developed an epitope-dependent agglutination assay to detect epitope-specific antibody response. Specifically, latex beads were coated with streptavidin and conjugated to one or more biotinylated epitope peptides. Antibodies specific to the epitopes were found to induce the agglutination of the corresponding latex beads in minutes (Fig. 5A), with the area of agglutination serving as a proxy of antibody titer. In principle, the latex bead agglutination assay detects the total antibodies (including IgG, IgM and IgA) rather than a specific isotype. To develop an epitope test to replace the S and N antigens, we coated the latex beads with the most prominent S or N epitopes. Specifically, latex beads were coated with a mixture of the S-811 and S-1146 (2S) peptides to represent the S antigen or the N-156 and N-361 (2N) peptides to represent the N antigen. When evaluated using plasma samples from individuals who tested positive (COVID+) or negative (COVID-) for the SARS-CoV-2 virus or samples from healthy donors collected in 2018 (PreCOVID), the 2S- and 2N-based agglutination assays showed 100% specificity and 99%-100% sensitivity (Fig. 5B).

**Figure 5.**
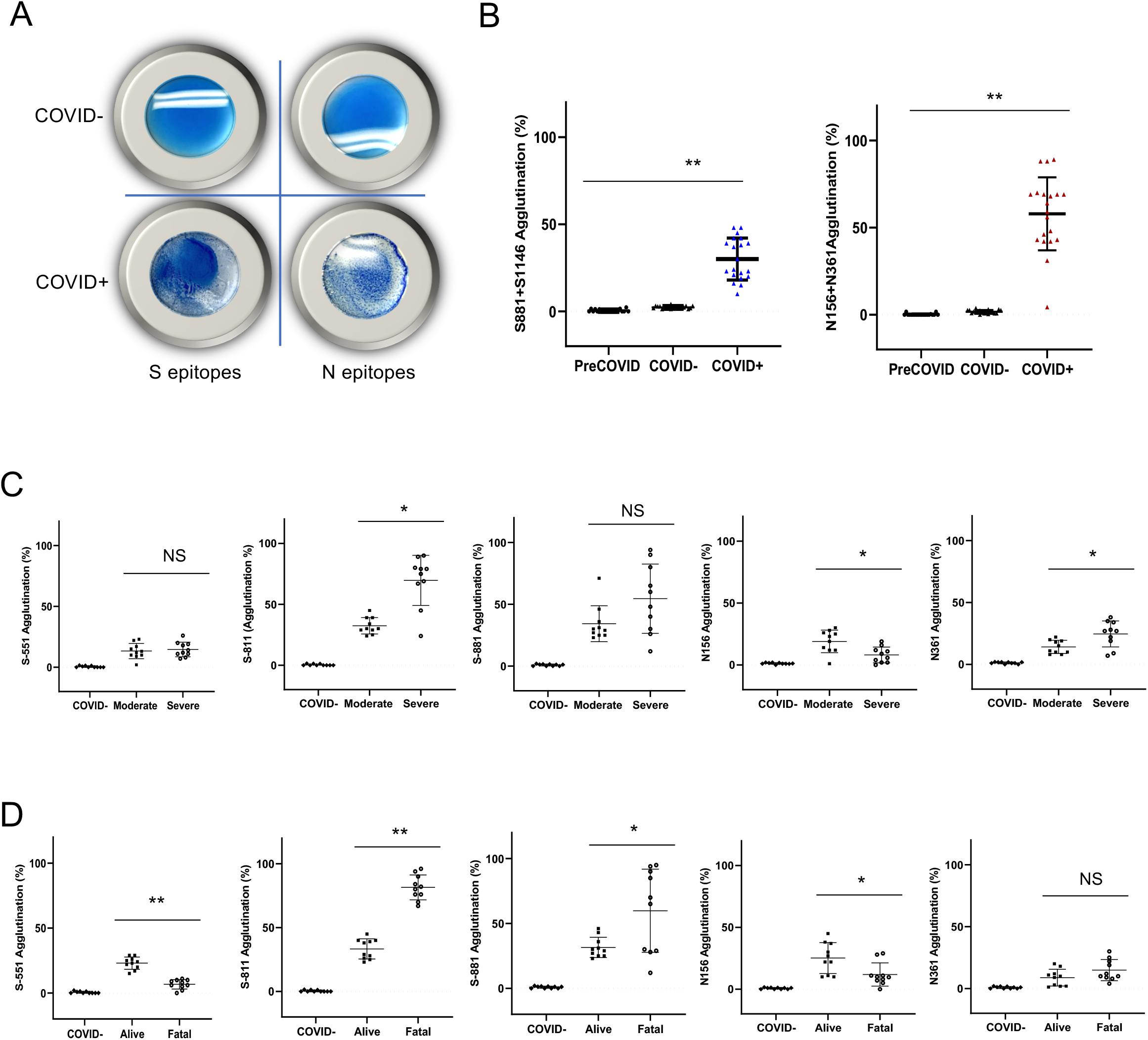
Rapid epitope-dependent agglutination assay for SARS-CoV-2 antibodies effectively differentiate patient groups. (**A**) Latex bead agglutination assay to gauge antibody response to SARS-CoV-2. The latex beads were coated with one or more biotinylated S or N epitope peptides and mixed with SARS-CoV-2 -negative (COVID-, top) or -positive (COVID+, bottom) plasma. The presence of antibodies against the epitopes promoted the agglutination of the latex beads. Images shown were taken after 2 min incubation at room temperature. (**B**) Epitope-based latex agglutination assay distinguished COVID-19+ from COVID-19- or preCOVID-19 plasma. The epitope peptides used were: S-811 and S-1146 from the spike and N-156 and N-361 from the nucleocapsid protein. (**C**) Correlation of disease severity with antibody responses to the S-811, N-156 and N-361 determined by latex bead agglutination. (**D**) Correlation of disease outcome with antibody responses to the S-551, S-811, S-881 and N-156 epitopes determined by latex bead agglutination. P values calculated based on unpaired Student t-test with Welch’s correction (no assumption of equal SD) (n=20 for B; n=10 for C & D). *p<0.05, **p<0.002.

To determine if the epitope-dependent agglutination assay could differentiate the different patient groups as effectively as the master epitope array, we coated the latex beads with the S epitope S-811, S-881 or S-551 or the N epitope N-156 or N-361 and performed agglutination assay on COVID-19 patient plasma or control (COVID-) specimen. While no agglutination was observed for the COVID-plasma, the COVID+ plasma promoted the agglutination of the latex beads in an epitope-dependent manner. We found that the group with severe disease had significantly greater S-811- and N-361-specific antibody responses than the group with moderate conditions. The reverse was found true for the N-156 epitope. Similarly, significant differences in antibodies specific for the S-811, S-881, S-551 and N-156 epitopes were observed between the alive and fatality groups. Notably, a high level of S-811-dependent agglutination was strongly and significantly correlated with patient death whereas even a moderate level of S-551-specific antibody response was correlated significantly with favorable outcome. These data not only reinforced our findings from the master epitope peptide array screen but extended these findings by identifying a group of key epitopes, including S-811, S-881, S-551 and N156, that collectively may help predict the clinical severity and outcome of the COVID-19 disease.

### Correlation of epitope-specific antibody response with neutralizing efficiency and disease outcome

Because neutralizing antibodies play a pivotal role in the humoral immune response to the SARS-CoV-2 infection, we used a surrogate neutralization assay to measure efficacy of patient plasma in blocking S-RBD binding to its host receptor, angiotensin converting enzyme 2 (ACE2) in vitro(29). We found that the neutralization efficiency of the plasma in the severe patient group was significantly higher than the group with moderate disease. Intriguingly, the plasma from the fatality group were significantly less efficient in neutralizing S-RBD binding to ACE2 compared to patients who recovered from the infection (Fig. 6A). This suggests that the ability to inhibit the S-RBD-ACE2 interaction, the critical first step in SARS-CoV-2 infection of host cells, dictates disease outcome. Because the identified S epitopes reside outside of the RBD domain of the spike protein, due perhaps to the possibility that the antibody-RBD recognition involves primarily conformational epitopes(23), we replaced the S epitopes with recombinant RBD and repeated the agglutination assay using the same plasma samples. We found that the S-RBD-dependent antibody response measured by latex agglutination significantly correlated with favorable outcome (Fig. 6B).

**Figure 6.**
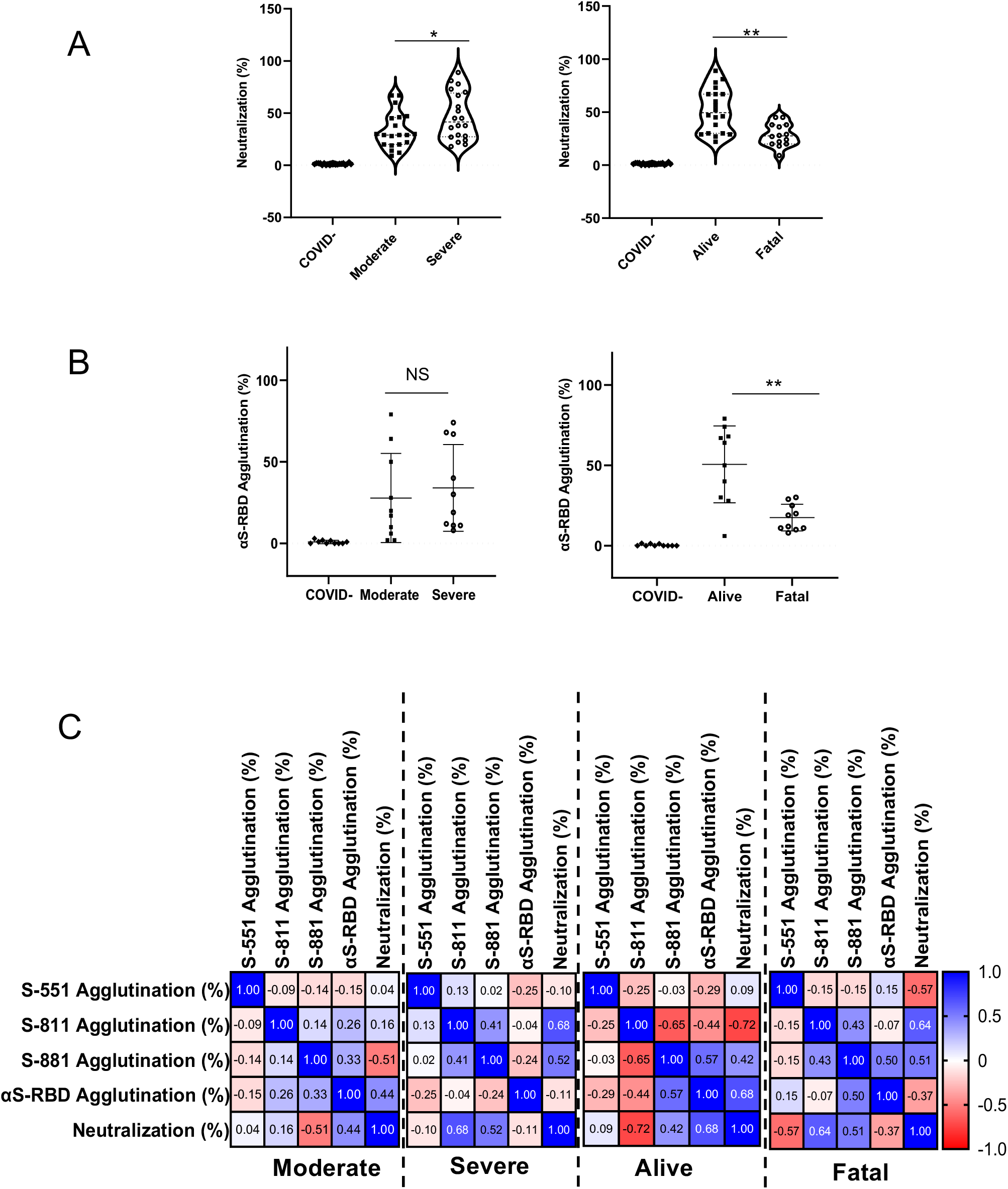
Antibody specificity predicts neutralization efficiency and disease outcome. (**A**) Correlation of neutralization efficiency with clinical severity (left) or outcome (right). *, p<0.05; **, p<0.01. (**B**) Correlation of S-RBD-antibody response measured by latex agglutination with COVID-19 severity (left) or outcome (right). **, p<0.01. (**C**) Pearson (r) correlation between epitope-dependent agglutination and neutralization. Confidence interval (CI): 95%. P values were based on one-way ANOVA with Geisser-Green house correction (no assumption of equal variability of difference) (n=20 for A, except n=14 for fatal; n=10 for B &C).

Can the epitope-specific antibody response be used to predict neutralization efficiency? We investigated this possibility by correlating the epitope-specific agglutination data with the neutralization data for the same set of patient samples. We found that the S-811-specific antibody response correlated with the neutralization efficiency negatively (r=-0.72, p<0.05) in the alive patient group, but positively in the severe (r=0.68) or fatality (r=0.64) patient group. In contrast, the S-RBD-dependent agglutination efficiency was positively correlated with neutralization efficiency (r=0.68) in the alive group(30) (Fig. 6C). Collectively, these data suggests that a strong S-RBD antibody response together with a weak S-811-specific response are indicative of favorable clinical outcome.

## DISCUSSION

The relationship between COVID-19 clinical severity and the humoral immune response is a complex one. It remains poorly understood to date why patients with severe symptoms are characterized with a stronger antibody response, including neutralization antibodies, to SARS-CoV-2 than those who have moderate or mild symptoms(30, 31). This dichotomy suggests that not all antibodies are beneficial. Indeed, while antibodies may mediate the clearance of the virus and virus-infected cells through antibody-dependent cellular cytotoxicity (ADCC) and phagocytosis (ADCP), they have also been proposed to play a pathogenic role via antibody-dependent enhancement (ADE)(32). The challenge is how to identify and differentiate between “good” and “bad” antibodies. Our epitope-based antibody analysis showed that the antibody responses from different patients are highly varied, and that there is generally no apparent association between the severity of disease presentation and antibody response measured using a protein antigen, including the Spike (N) or Nucleocapsid (N). Therefore, antibody profiling with greater resolution than a simplified S or N antibody classification is needed. Our work, which combines both systematic antibody screen using peptide/protein arrays and rapid antibody assays based on latex agglutination, showed that epitope-resolved antibody testing is far more sensitive than S/N-based serology tests in discerning antibody specificity and identifying the correlates between humoral immunity and COVID-19 disease severity or patient survival.

By identifying and validating the major S and N epitopes to enable epitope-specific antibody testing, our study not only greatly extended previous work that show linear epitopes play a critical role in mediating antibody responses to SARS-CoV-2 (19, 20, 22, 24, 33), but more importantly, it demonstrated that the complex antibody responses in individual patients may be deconvoluted by epitope-resolved antibody profiling. Systematic and unbiased antibody profiling using a master array comprising the most prominent epitopes led to several intriguing findings. First, patients with severe disease or poor outcome tend to have antibodies against a large number of epitopes. We showed that these same patients had low levels of neutralizing antibodies. It is therefore likely that the increased production of non-neutralizing antibodies contributed to disease development. Second, all epitopes are not equal, and even the epitopes from the same protein antigen (S or N) may play distinct roles in dictating disease severity and outcome. We have shown that not only S-811 and S-881 are two of the most prevalent epitopes, but also a high level of antibodies specific for these epitopes are strongly indicative of severe or fatal disease. That the S-811 antibody response is negatively correlated with neutralization efficiency in patients who survived the infection suggests that the S-811-driven antibody response may be indicative of more aggressive or effective virus infection of the host cells. Alternatively, antibody response targeting these epitopes may be a surrogate marker for a more robust and potentially excessive immune response causing greater tissue injury. Third, we have shown that mutations found in SARS-CoV-2 variants may directly affect antibody response by altering epitope specificity. This finding demonstrated the flexibility of the epitope peptide array approach to quickly incorporate emerging mutations, thereby providing valuable information of the mutations and the corresponding variants on the immune system.

While it has been shown that mutations result in more fit, and likely more contagious viruses(15, 16, 34), the serological consequences of the mutations found in SARS-CoV-2 variants are unclear(34). Recent studies have shown reduced binding to therapeutic antibodies or Spike-specific antibodies for the circulating variants B.1.1.7, 501Y.V2 and P.1 in vitro (35, 36), suggesting that mutations in these variants directly affect antibody response. In agreement with this assertion, we found that certain mutations, including the P681H in the Spike and S245F in the Nucleocapsid proteins, rendered the corresponding epitopes completely incapable of binding antibodies specific for the original virus. This observation raises concerns over the effectiveness of vaccines in protecting against the coronavirus variants that are currently circulating or variants, including variant recombination, that may emerge in the future (14). While it remains to be determined whether this mutation could mediate immune escape of the variant in some patients, our finding may have far-reaching implications as it may render the wild-type Spike mRNA-based vaccine less effective to those who employ S-671 (which encompass the mutated residue) as a major epitope. However, we note that the P681H and S235F mutations only affected a few individuals in the cohort of patients examined herein while the majority of patients displayed no apparent antibody responses against the corresponding epitopes. This may explain why recent studies have shown the Pfizer and Moderna mRNA vaccines are effective in protecting from infection by the variant (37-39). It would be important to investigate in the future, by large-scale epitope-specific antibody profiling, the percentage of the population who employ S-671 as a major epitope. By the same token, prevalence of the N-221 epitope (which contains the S235 residue found mutated in the UK strain) would provide valuable information on the protection of vaccines based on inactivated intact viruses. In the same vein, hundreds of mutations may be examined simultaneously in a peptide array to assess their effect on antibody response, and the epitope array may be readily modified to incorporate emerging mutations. The impact of the mutations on humoral immune response may also involve conformational epitopes which are not recapitulated by the linear epitopes. However, both the master array and the agglutination-based antibody test may be quickly modified to include emerging mutations.

Future studies using a combination of epitope and protein antigen-based assays tailored to emerging variants would provide valuable information on the population penetrance of a given variant and the impact the associated missense or deletion mutations on antibody-mediated immunity.

While the epitope array may be used to profile antibody response in a systematic manner, the epitope-dependent latex agglutination assay provides a rapid, simple, cost-effective, and accurate serological test that may be suitable for POC antibody testing. The agglutination assay may be carried out with individual epitopes to map the specificity of antibodies or with a mixture of epitopes to test multiple antigens simultaneously. The ease with which to incorporate mutated epitopes or S/N protein antigen in the agglutination assay makes it a nimble yet powerful tool to determine the impact of mutations associated with the coronavirus variants on humoral immunity. Although the mRNA-based vaccines have shown superb efficacy, not all vaccine recipients would be protected. It also remains to be determined how long the immunity will last and against which variant. Monitoring vaccinated or recovered individuals over months to years by antibody testing would provide valuable information on the duration of immune responses against SARS-CoV-2, including variants (7). In this regard, the epitope-resolved antibody test may be used to delineate the specific antibodies produced by different individuals, determine persistence of antibody in the circulation over time, assess the efficiency of vaccines, and decipher the effect of emerging variants on the immune system. The agglutination assay, which measures the total antibody response irrespective of the Ig isotypes, provides a unique advantage over serological assays that measure a given isotype as different Ig isotypes have distinct dynamics and evolutionary trajectory over time(18). Longitudinal studies by the epitope-resolved agglutination assay would provide valuable information on the evolution of antibody immunity from vaccination or previous infection.

With the rapid spread of the highly contagious coronavirus variants, the number of patients that need treatment will likely remain high for quite some time. In the absence of an effective treatment, convalescent plasma and antibody therapy remain a good option. However, clinical trial results are mixed for either approach, highlighting the need for more specific screening strategies to identify the most effective convalescent plasma or therapeutic antibody. It would be interesting to find out if the epitope-resolved antibody profiling strategy developed herein may be used to identify convalescent plasma donors with a high level of neutralization antibody and a low level of non-neutralizing and potentially harmful antibodies. Similarly, the epitope specificity mapping could enable the selection of the most effective therapeutic antibody with the least side-effects.

## METHODS

### Blood sample collection

Blood samples were collected following a protocol (study number: 116284) approved by the Research Ethics Board (REB) of Western University. The residual plasma samples were de-identified prior to transfer from the Core Laboratory (London Health Sciences Center, London, Canada) to a biosafety Level 3 (CL3) lab (ImPaKT, Western University) following Transportation of Dangerous Goods (TDG) guidelines. All plasma samples were heat-inactivated at 56 °C for 30 minutes at the ImPaKT CL3 facility as per Western University biosafety regulation. Heat inactivated plasma samples were then transferred to the testing laboratory.

### Protein Microarray

#### Proteins

The Spike-ectodomain(40), Nucleocapsid-dimerization domain, Nucleocapsid-RNA binding domain, NSP3-unique, NSP3-ADRP, NSP3-NAB, NSP3-PLPro, NSP4-CTD, NSP5, NSP7, NSP8, NSP9, NSP10, NSP16(41) were supplied by the Toronto Open Access Covid-19 Protein Manufacturing Center (comprising BioZone and the Structural Genomics Consortium (SGC)) under an Open Science Trust Agreement http://www.thesgc.org/click-trust. The Center received funding from the Toronto COVID-19 Action Fund. See Supplementary Table S1 for a complete list of proteins

#### Protein Array Printing

SARS-CoV-2 proteins were diluted to 0.5-10μM in PBS with 5% glycerol (IgG control at 200nM) and aliquots transferred to a 384-well microplate (ArrayIt). 24 copies of the microarray were printed on each nitrocellulose coated glass slide (ArrayIt) using a VersArray Chipwriter Pro (Bio-Rad) equipped with a Stealth 15XB microarray quill pin (ArrayIt). Spot to spot distance was 850μm with two reprints of the same spot and all spots printed in duplicate in the y dimension. A dwell time of 0.1sec was used for each spot with an approach speed of 12.5mm/sec. Samples were printed at room temperature and subsequently stored at 4°C until time of probing.

### Peptide Microarray

#### Peptide synthesis

Peptides were synthesized on Tentagel resin on an Intavis MultiPep RSi peptide synthesizer using N-(9-fluorenyl) methoxycarbonyl (Fmoc) chemistry. All peptides were synthesized with biotin at the N terminus followed by an aminohexanoic acid and Gly-Gly spacer. A walking array of peptides with 15 amino acid length and 5 amino acid overlap spanning the full sequence of SARS-CoV-2 Spike and Nucleocapsid proteins were synthesized for array printing. Peptides reported in a previous publication(24) as well as epitopes predicted using bioinformatics(25) were synthesized and printed to create the literature-reported peptide array. Peptides encompassing mutation sites reported in SARS-CoV-2 variants were synthesized as described above for the variant peptide array (Table 2).

#### Peptide Array Printing

Peptides were printed as neutravidin complexes on nitrocellulose coated slides (ArrayIt) by mixing 10μM neutravidin with an excess (by four-fold) of peptide that was diluted in phosphate buffered saline and aliquots transferred to a 384-well microplate (ArrayIt) along with IgG printing control, Spike RBD, full length Nucleocapsid, N-RBD and N-Dimerization proteins. 2 copies of the walking microarray, 3 copies of the literature-reported microarray, or 8 copies of the variant array were printed on each nitrocellulose coated glass slide using a VersArray Chipwriter Pro (Bio-Rad) equipped with a Stealth 15XB microarray quill pin (ArrayIt). Spot to spot distance was 750μm with two reprints of the same spot and all spots printed in duplicate in the y dimension. A dwell time of 0.1sec was used for each spot with an approach speed of 12.5mm/sec. Samples were printed at room temperature and subsequently stored at 4°C until time of probing

### Protein and Peptide Array Probing

Microarray slides were briefly rinsed twice with TBST (Tris buffered saline containing Tween 20: 0.1M Tris-HCl, pH 7.4, 150mM NaCl, and 0.1% Tween 20) to wet the surface and then incubated for 2 hours with ChonBlock ELISA blocking and antibody dilution buffer (Chondrex Inc). Slides were briefly rinsed with TBST then inserted into an ArraySlide 24-chamber hybridization cassette (The Gel Company) for the proteome array or ProPlate Multi-Well Chamber (Grace Bio-Labs) for the peptide arrays and incubated with plasma from SARS CoV-2 NAT confirmed positive and negative patients (1:250 dilution in ChonBlock). Slides were then rinsed quickly three times followed by three 5min washes with TBST before probing with goat anti-human IgG HRP antibody at 1:10,000 (Millipore Sigma) in ChonBlock for 1h. The wash step was repeated as above, then the HRP signal was visualized on a ChemiDoc XRS+ Imager (Bio-Rad) using Clarity ECL Substrate (Bio-Rad). Slides were incubated with ECL solution for 30 seconds then 15 images were taken incrementally from 1-60 seconds. All incubation steps were performed at room temperature using a rocker for agitation of sample.

### Array Quantification

Peptide walking arrays, literature-reported epitope peptide arrays and the master epitope arrays were quantified using ImageJ software(42). Images were first inverted and converted to 8-bit. Background was subtracted using a rolling ball radius of 25.0 pixels. Intensities were normalized to IgG control and ranked by normalized signal intensity. Peptides with strongest intensity or most frequently observed were selected for creation of the master array. For the master array, samples with no detectable antibody response or samples with signals within 2 standard deviations of the mean background intensity were omitted from statistical analysis.

### Preparation of SARS-CoV-2 peptide antigen conjugated latex particles and peptide antigen-based agglutination assay

Blue dyed carboxylate-modified, streptavidin-polystyrene latex beads, 0.25 µm in diameter or blue dyed polystyrene latex beads, 0.8 μm in diameter, were purchased from Sigma Aldrich (L6155, L1398). Carboxylate-modified latex-streptavidin or neutravidin coated polystyrene beads were suspended at 2.5% (w/v) using assay buffer, 0.025M MES-Tween 20 buffer (2-(N-Morpholino) ethanesulfonic acid, 0.05% pH 6.0). Synthetic biotin-labeled SARS-CoV-2 peptides were suspended in the same assay buffer at the concentration 500µg/ml. The biotin-peptides were incubated with streptavidin-latex beads for 1 hour at room temperature. The epitope peptide-conjugated latex beads complex was washed twice with PBS buffer (135 mM NaCl, 2.6 mM KCl, 8 mM Na2HPO4, and 1.5 mM KH2PO4, pH 7.4) by mixing and centrifuging the latex suspension at 5,000g for 10 min. The peptide antigen-bead conjugate was blocked for 30 min at room temperature in PBS containing 3% bovine serum albumin (BSA). The conjugate was then resuspended at 2.5% (w/v) in PBS containing 1% BSA and stored at 4°C until use. For the agglutination assay, 5 µl plasma was mixed with 25 µl peptide-conjugated latex beads (2.5%, w/v) per assay as described in the full protein antigen agglutination assay.

### Agglutination assay for SARS-CoV-2 antibody testing and data interpretation

For the agglutination assay, 5 μl plasma was mixed with 25 µl antigen-coated beads (2.5%, w/v) per assay. The agglutination was allowed to proceed for 2 min at room temperature before imaging with a camera. The relative degree of agglutination induced by the SARS-CoV-2 antibody was measured by the area of clump formation based on the corresponding image. The image analysis software Qupath (v0.1.2) was used (https://qupath.github.io/) and quantification was done by calculating the percentage of agglutination based on estimated agglutination/clumps area (mm^2^) relative to the total latex reaction area.

### S-RBD-ACE2 binding ELISA surrogate neutralization assay

Biotin-ACE2 (1µg/m) was added to S-RBD-coated plate after blocking and incubated for 1hour at room temperature. The wells were washed 3 times with TBST (20 mM Tris, 150 mM NaCl, 0.1% Tween 20) to remove unbound biotin-ACE2. Streptavidin-HRP (1000-fold dilution with Chonblock blocking buffer) was then added to each well and incubated for 1hour at room temperature. The wells were washed 3 times with TBST and TMB substrate (3,3’,5,5’-Tetramethylbenzidine, Thermo Scientific, N301) was added for reaction development and 0.18 M H2SO4 was used to stop reaction. Absorbance at 450nm was measured to detect the S-RBD bound ACE2. To determine the neutralization efficacy of the patient plasma, the plasma was diluted 1:100 and incubated with S-RBD-coated wells (blocked) for 1hour at room temperature. The wells were washed three times with TBST. Biotin-ACE2 was then added to the wells and incubated for 1 hour at room temperature followed by washing, reaction development and detection as described above.

### Statistical analysis

All statistical analyses were done using the GraphPad Prism9 software.

## Data Availability

Supplementary data available upon request

## AUTHOR CONTRIBUTIONS

SSCL, ICY, CV, SE and XL conceived and designed the study. CV performed the protein and peptide array screens and data analysis. SE performed the agglutination and neutralization assay and data analysis. XL, SZ synthesized the peptides. SA, PS, AS, AH, FY, TS, EE, PL, PG, and TH contributed to subcloning, expression and purification of SARS-CoV-2 proteins and human ACE2. MK, MJK, LL, HA, BDH, VB, CMM, MS, DF contributed to patient recruitment, blood sample collection and NAT and ELISA testing. TK and BCY contributed to data analysis and interpretation. SSCL, CV and SE wrote the manuscript with input from ICY, BCY, SA, MK, and DF.

## ACKNOWLEDGEMENT

We thank the Center for Structural Genomics of Infectious Diseases (CDGID) for the N-RBD expression plasmid, Dr. Jason McLellan for human ACE2 and Spike ectodomain expression plasmids, Dr. Shane Harding for the spike-receptor binding domain expression construct, and Yanjun Li for technical assistance. This work was supported by grants from the Ontario Research Fund-COVID-19 Rapid Research Fund, the Toronto COVID-19 Action Fund, Western University (Research), the Departments of Medicine and Pediatrics at Western University, the Lawson Health Research Institute (https://www.lawsonresearch.ca/), the London Health Sciences Foundation (https://lhsf.ca/), and the AMOSO Innovation Fund. SE was supported by a Post-Doctoral Fellowship from the National Science and Engineering Council of Canada. SSCL holds a Canada Research Chair in Molecular and Epigenetic Basis of Cancer.

